# ANTIBODY RESPONSE TO COVID-19 INFECTION- CLINICAL VARIABLES AT PLAY

**DOI:** 10.1101/2020.11.20.20234500

**Authors:** Anuj Parkash, Parul Singla, Meenu Bhatia

## Abstract

**Background:** The current COVID19 pandemic began in December 2019 and rapidly expanded to become a global pandemic. The COVID 19 presents multitude of clinical disorders, ranges from asymptomatic infection to severe disease, which can accompanied by multisystem failure leading to death. The immune response to SARS CoV 2 is understood to involve all the components of the system that together causes viral elimination and recovery from the infection. However, such immune responses implicated in the disease has varied presentation ranging from mild to a severe form, which appears to hinge on the loss of the immune regulation between protective and altered responses. In this study, we want to unravel this association of immune responses to various clinical variables, which might have a major role to play, while generating the immune response. The objective was to test this hypothesis in our settings and comparing the results of serologic tests from a group of COVID 19 patients and will analyzed the disease severity in comparison.

**Methods:** Testing for SARS COV2 IgG Antibody was done with chemiluminescent assay on the Ortho Clinical Diagnostic’s (OCD) Vitros 5600 platform.

**Results:** A total of 106 COVID 19 patients were included in this study, of whom 61 were male and 45 were female. Their mean age was 43.7 years (range 17–83) and the median interval between initial symptom onset and sample collection was 12.33 days. Eighty patients (82%) had mild or moderate symptoms and twenty-six patients (18%) had severe symptoms. The antibody titers were positive in 99 patients (93%) and were found negative in 7 patients (7%). When comparing patients with mild/moderate symptoms and patients with severe/critical diseases, no statistically significant difference was observed between their gender ratios (P = 0.373) and age composition (P = 0.224).

**Conclusions:** The data presented in this research study did not find any statistical significance between SARS CoV 2 IgG antibody levels with COVID 19 disease severity, duration of symptoms, age, gender, and length of convalescence.

## INTRODUCTION

The ongoing pandemic of COVID 19 is causing big threat to communities around the world. It is caused by a virus called severe acute respiratory syndrome coronavirus 2 (SARS-CoV-2). It is a highly contagious virus and spread by respiratory route. Patients most commonly presented with fever, cough and respiratory symptoms. Mortality and morbidity quite largely depends upon the host immune response and people with co-morbidities are at greater risk from COVID-19 infection.. As a general immune principle, body immune response to acute phase of infection starts with IgM antibody, whereas IgG and IgA antibodies appears 3 to 4 weeks to post-exposure ^1,2^. However, it has been observed that the immune response against this virus is highly variable in nature. It is a general assumption that the patients who have had more severe disease appear to have higher levels of important neutralizing antibodies^3^. Similarly, patients who had mild or asymptomatic COVID-19 have low levels of neutralizing antibodies. Many authors have reported diverse presentation of SARS-CoV-2 antibodies and varied presentation with different clinical features ^4,5,6,7^. The present study is undertaken to understand the status of antibody generation in confirmed COVID-19 patients and to analyze its correlation with disease severity, if any in our hospital settings.

## Material and Method

The present study is a single-center, open-label, cross-sectional, retrospective study. The Subjects were recruited into single-arm consulting COVID19 OPD of a tertiary care cancer hospital in North India. All the subjects presenting to our hospital from the start of the pandemic to 11 September 2020, with COVID-19 infection were eligible for enrolment into the study. In keeping with Indian Council of Medical Research (ICMR) protocols and international practice, patients were deemed to have COVID-19 if a Real-time Polymerase chain reaction (RT-PCR) assay test and/or cartridge-based nucleic acid amplification test (CBNAAT) test from a combination of the naso-oropharyngeal swab was positive for SARS-CoV-2. Patients with a radiological or clinical suspicion of COVID-19, but a negative RT-PCR test were not included in this analysis. A total of 108 subjects were enrolled in the study who were tested positive either by RT PCR) or CBNAAT by GeneXpert. The testing data were extracted from hospital electronic medical records (EMR) and the clinical data was obtained telephonically for detailed medical history. The data set includes patient demographics, co-morbidities, and course of illness, various symptoms, type of symptoms, and the severity of symptoms specific to COVID-19.

### Method

Testing for SARS COV2 IgG Antibody was done with chemiluminescent assay on the Ortho Clinical Diagnostic’s (OCD) Vitros 5600 platform, US with the dedicated reagents. Antibody levels are presented as chemiluminescence values divided by cutoff (S/CO) levels. The cutoff level used in the study was 1.00, as per the kit insert from OCD. The RT PCR test was performed on Quant 5 studio (Thermo-Fisher life science) employing an ICMR approved real-time test kit (TRUPCR®), whereas CBNAAT testing was done with Cepheid®USA (GeneXpert® assay).

### Statistical Analysis

Continuous variables are presented as Mean±S.D, whereas for categorical variables, frequencies, and percentages were provided. Shiparo-Wilk test was used to test the normality of the data for the data which follows the normal distribution (P-value >0.05). An Independent t-test was used to compare the age and duration of symptoms between Mild/Moderate and Severe patients. A Chi-square test was used to compare categorical variables with the severity of symptoms. All data entries and statistical analyses have been performed by using SPSS® Version 23.0 software. All the reported p-values were two-sided and p-values <0.05 were considered to indicate statistical significance.

### Ethical approval

The study was approved by the Institution Review Board (IRB) (ref. no. 2020003). The written informed consent was waived by the IRB, since the study was retrospective in nature.

## Results

A total of 106 COVID-19 patients were included in this study, of whom 61 were male and 45 were female. Their mean age was 43.7 years (range 17–83) and the median interval between initial symptom onset and sample collection was 12.33 days. Eighty patients (82%) had mild or moderate symptoms and twenty-six patients (18%) had severe symptoms. (Table 1). The antibody titers were positive in 99 patients (93%) and were found negative in 7 patients (7%). When comparing patients with mild/moderate symptoms and patients with severe/critical diseases, no statistically significant difference was observed between their gender ratios (*P* = 0.373) and age composition (*P* = 0.224). The average duration of the convalescence period was 12.14 days to 12.92 days in mild to moderate cases and severe cases respectively with no statistical significance. Also, while analyzing various symptoms of the patients, the majority of patients experienced fever (97%), myalgia (61%) cough (60%), and weakness (37%) (Table 2). However, few patients experienced other symptoms e.g. loss of taste, loss of smell, diarrhea, and gastritis with minor frequency. The antibodies were detectable in samples collected over 90 days after onset of disease with the earliest seroconversion of IgG antibody was observed 2 days after disease onset. The area under the curve (AUC) of receiver operating characteristic curve (ROC) to predict the best possible detection of antibodies in the serum post-infection was 0.704 with 95% CI (0.603-0.792) (Fig 1).

**Table 1:** Demographic information

| Severity of Symptoms | Mild/Moderate | Severe | P-value |
| --- | --- | --- | --- |
| Total | 80 | 26 | N/A |
| Age | 42.55±16.16 | 47.12±16.46 | 0.224 |
| Duration of convalescence | 12.14±1.78 | 12.92±2.88 | 0.100 |
| Female | 36 | 9 | 0.373 |
| Male | 44 | 17 |  |
| Negative | 5 | 2 | 0.797 |
| Positive | 75 | 24 |  |

**Table 2:** Type of symptoms and their distribution

| Variables |  | Mean±S.D/Frequency(%) |
| --- | --- | --- |
| Age |  | 43.67±16.27 |
| Duration of symptoms |  | 12.33±2.11 |
| Sex | Female | 45(42.5) |
|  | Male | 61(57.5) |
| SARS COV 2 IgG Antibody | Negative | 7(6.6) |
|  | Positive | 99(93.4) |
| Fever | Absent | 3(2.8) |
|  | Present | 103(97.2) |
| Weakness | Absent | 66(62.3) |
|  | Present | 40(37.7) |
| Cough | Absent | 46(43.4) |
|  | Present | 60(56.6) |
| Headache | Absent | 81(76.4) |
|  | Present | 25(23.6) |
| Dyspnoea | Absent | 98(92.5) |
|  | Present | 8(7.5) |
| Pneumonia | Absent | 105(99.1) |
|  | Present | 1(0.9) |
| Gastritis | Absent | 93(87.7) |
|  | Present | 13(12.3) |
| Myalgia | Absent | 41(38.7) |
|  | Present | 65(61.3) |
| Nasal Congestion | Absent | 101(95.3) |
|  | Present | 5(4.7) |
| Loss of taste | Absent | 103(97.2) |
|  | Present | 3(2.8) |
| Loss of smell | Absent | 102(96.2) |
|  | Present | 4(3.8) |
| Diarrhoea | Absent | 100(94.3) |
|  | Present | 6(5.7) |
| Vomiting | Absent | 105(99.1) |
|  | Present | 1(0.9) |

**Figure 1:**
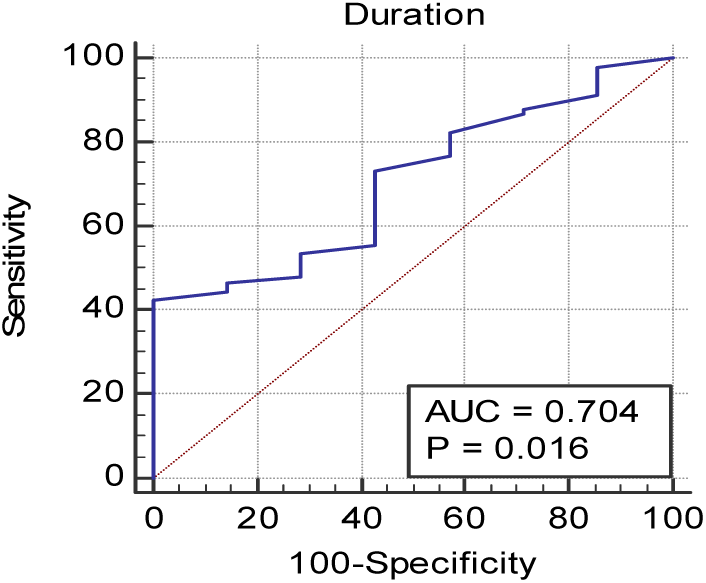
AUC of receiver operating characteristic curve (ROC) to predict the best possible detection of antibodies.

## DISCUSSION

COVID□19 represents an array of various clinical forms ranging from asymptomatic to mild, moderate, and very severe cases. In the present study out of 106 patients, males are affected more than females. This varied presentation could be due to higher exposure risk to males. Around 82% patients present with mild to moderate symptoms and 18 % patients present with severe symptoms. Majority of subjects (97.2%) present with fever and 40-50% subjects complaint of associated symptoms of cough, myalgia and weakness. More than 95% cases did not give history of any loss of taste or smell or Nasal congestion. Sharma et al in the review article on global trends of clinical presentation also enumerated similar trends of clinical presentation.^8^ The immunology of all these cases is different and becomes more complex when the patients progresses to severe disease. The research studies done by Long et al, Thevarajan et al, have found an association between high SARS-CoV-2 antibody levels and COVID-19 severity ^3, 9^. A plausible explanation for this observation may be that patients experiencing severe forms of the disease are exposed to higher viral load ^9^. Alternatively, it may represent an overall exaggerated immune response driven either by “cytokine storms” or by “antibody-dependent enhancement” (ADE) which are two known immune responses causing severe lung tissue damage. However, as a counter explanation, many authors have found no association of the antibody response against COVID-19 with the severity of the disease. A recent study by Phipps W et al^10^, has concluded that the measurement of IgG and IgM antibodies did not predict disease severity. The data studied herein also do not support the abovementioned association between high SARS-CoV-2 antibody levels and COVID-19 severity. In effect, we failed to find differences in SARS-CoV-2 IgGs levels between patients with mild, moderate, or severe symptoms, when they are matched for age, sex, and co-morbidities. The threshold duration of antibody detection was 7 days from the onset of disease with sensitivity and specificity of 71.11 and 57.14 respectively with area under the curve (AUC) of 0.704 with 95% CI (0.603-0.792). Similar findings regarding antibody detection after COVID-19 illness has been observed in other clinical studies by Rode etal^11^. Antibody dynamics due to variable antigen presentation may contribute to false positive or negative results and inconsistencies in seroprevalence studies^12^. In our opinion, a comparison between studies addressing the abovementioned issue is challenging, thanks to notable variations in clinical characteristics, categorization of severity, and ways SARS-CoV-2 antibodies are utilized for detection and quantification.

## CONCLUSION

The data presented in this research study did not find any statistical significance between SARS-CoV-2 IgG antibody levels with COVID-19 disease severity, duration of symptoms, age, gender, and length of convalescence. We finally conclude that a better understanding of the pathogenesis and immunogenicity should be studied in greater detail to understand the ever changing immunogenicity profile of SARS-COV-2.

## Data Availability

The raw data is available with the author and can be reproduced on demand.

## ACKNOWLEDGEMENT

We would like to acknowledge Dr Anurag Mehta for his guidance and our patients who helped us in conducting the study.

